# Pharmacokinetics and safety of XAV-19, a swine glyco-humanized polyclonal anti-SARS-CoV-2 antibody, for COVID-19-related moderate pneumonia: a randomized, double-blind, placebo-controlled, phase IIa study

**DOI:** 10.1101/2021.04.15.21255549

**Authors:** Benjamin Gaborit, Eric Dailly, Bernard Vanhove, Régis Josien, Karine Lacombe, Vincent Dubee, Virginie Ferre, Sophie Brouard, Florence Ader, Marie-Anne Vibet, Aurélie Le Thuaut, Richard Danger, Laurent Flet, Anne Omnes, Laetitia Berly, Anne Chiffoleau, Alexandra Jobert, Odile Duvaux, François Raffi, POLYCOR trial group

## Abstract

**Background:** We assessed the pharmacokinetics and safety of XAV-19, a swine glyco-humanized polyclonal antibody against SARS-CoV-2, in COVID-19-related moderate pneumonia. In vitro, 100% neutralization activity is seen with XAV-19 concentrations above 5 μg/mL.

**Methods:** In this phase 2a trial, adults with COVID-19-related moderate pneumonia of ≤10 days duration were randomized to infusion of XAV-19 0.5 mg/kg at day 1 and day 5 (group 1), 2 mg/kg at day 1 and day 5 (group 2), 2 mg/kg at day 1 (group 3) or placebo.

**Results:** Eighteen patients (n=7 for group 1, n=1 for group 2, n=5 for group 3, and n=5 for placebo) were enrolled. Baseline characteristics were similar across groups, XAV-19 serum concentrations (μg/mL, median, range) at C_max_ and at day 8 were 9.1 (5.2-18.1) and 6.4 (2.8-11.9), 71.5 and 47.2, and 50.4 (29.1-55.0) and 20.3 (12.0-22.7) for groups 1, 2 and 3, respectively (p=0.012). Terminal half-life (median, range) was estimated at 11.4 (5.5-13.9) days for 2 mg/kg of XAV-19 at day 1. Serum XAV-19 concentrations were above the target concentration of 10 μg/mL (tow fold the in vitro 100% inhibitory concentration [IC100]) from the end of perfusion to more than 8 days for XAV-19 2 mg/kg at day 1. No hypersensitivity or infusion-related reactions were reported during treatment, there was no discontinuation for adverse events and no serious adverse events related to study drug.

**Conclusions:** Single intravenous dose of 2 mg/kg of XAV-19 demonstrated high serum concentrations, predictive of potent durable neutralizing activity with good tolerability.

**Trial registration:** ClinicalTrials.gov Identifier: NCT04453384

**Main point:** In this first-in-human trial including patients with COVID-19-related pneumonia, a single 2mg/kg dose of a swine glyco-humanized polyclonal anti-SARS-CoV-2 antibody, achieved serum concentrations above the target of neutralization threshold for 8 days in all patients, with good tolerability and safety.

## INTRODUCTION

Since the first identification of severe acute respiratory syndrome coronavirus 2 (SARS-CoV-2) from patients with bilateral pneumonia, in Wuhan, China, during December 2019, the ongoing pandemic of coronavirus disease 2019 (COVID-19) has affected more than 127 Million people and caused death in 2.8 Million [1,2]. Among patients hospitalised for COVID-19, 15-20% develop a severe respiratory failure requiring admission to intensive care unit [3]. Corticosteroids and anticoagulant therapy have improved prognosis of patients requiring respiratory support for severe or critical pneumonia [4,5]. Multitargeted interventions in severe COVID-19, combining potent antiviral(s), steroids, anticoagulants and in most severe cases adjunctive immune-based therapy such as tocilizumab, could constitute an optimized cocktail to halt further progress to respiratory failure, acute respiratory distress syndrome (ARDS), multi-organ dysfunction, and death [6–8].

In an ongoing trial, we are investigating XAV-19, a swine glyco-humanized polyclonal SARS-CoV-2-neutralizing antibody [9], in hospitalized patients with COVID-19 pneumonia requiring low flow oxygen supplementation [10]. The main hypothesis is that reducing viral burden and improving specific immunity by passive antibodies administration at hospital entry of patients hospitalized for COVID-19-related moderate pneumonia within 10 days of first symptom onset could lead to clinical benefit. Animal-derived heterologous polyclonal antibodies, used as a passive heterologous immunotherapy, could represent a highly efficient alternative to the use of monoclonal antibodies in COVID-19 by targeting multiple antigen epitopes. The dose rationale for the administration of XAV-19 was based on assessment of in vitro inhibitory potency of XAV-19 on SARS-CoV-2 virus and the target serum concentration was established at 10 μg/mL [9,11].

Here, we report results of the first part of this ongoing clinical trial involving patients hospitalized for COVID-19-related moderate pneumonia requiring oxygen supplementation. In this phase IIa of the trial, objectives of this first-in-human administration of XAV-19 were to assess its pharmacokinetics and safety.

## METHODS

### Study design and participants

This is an ongoing multicenter, randomized, double-blind, placebo-controlled, phase IIa-III clinical trial involving hospitalized patients with COVID-19-related moderate pneumonia requiring low-flow oxygen supplementation[10]. The first part was conducted as a phase IIa, first-in-human dose-ranging study at four sites in France, to assess the pharmacokinetics and safety of XAV-19, to select the optimal dose for the second part of the trial, designed as a phase III for which recruitment is ongoing.

This study was conducted in accordance with good clinical practice procedures, all applicable regulatory requirements, and the guiding principles of the Declaration of Helsinki. The study protocol was reviewed and approved by the Ethics Committee OUEST VI (20.06.15.31306). All patients provided written informed consent before the entry in the study. The study is registered with ClinicalTrial.gov, NCT04453384.

We prospectively identified hospitalized adults aged between 18 and 85 years, with SARS-CoV-2 infection confirmed by a positive RT-PCR and onset of first symptoms less than 10 days prior to enrolment. Inclusion criteria included COVID-19-related moderate pneumonia defined by SpO2 ≥ 92% on oxygen supplementation ≤ 6L/min by low-flow nasal cannula or mask (score of 4 on the World Health Organization 7-point Clinical Progression Scale (WHO-CPS)) and evidence of pulmonary involvement on lung examination (rales/crackles) and/or chest-imaging (Chest X-ray or computed tomography). Exclusion criteria were evidence of multiorgan failure, receipt of immunoglobulins or any blood products in the past 30 days, psychiatric or cognitive illness or recreational drug/alcohol use that would affect subject safety or compliance, end-stage renal disease (eGFR < 15 mL/min/1,73 m^2^), child-Pugh C-stage liver cirrhosis, decompensated cardiac insufficiency, known allergy, hypersensitivity or intolerance to the study drug or to any of its components, life expectancy estimated to be less than 6 months patient under guardianship or trusteeship, pregnancy or lack of effective contraception in women of childbearing potential.

### Randomization and masking

Eligible participants were assigned to two consecutive groups of approximately eight patients per cohort of ascending dose, in a 1:1 randomization scheme per dose for the first two patients and a five active and one placebo randomization scheme per dose for the six following participants of each cohort. The two doses of XAV-19 were 0.5 mg/kg and 2 mg/kg administered over one hour intravenous infusion at days 1 and 5. After inclusion of the first two participants in each cohort, the safety and tolerability of the treatment was assessed up to 8 days post-infusion by an independent data monitoring committee before continuation of enrolment. Following blinded analysis of the first cohort (0.5mg/kg at D1 and D5) and of the first two patients (one active and one placebo) of the second cohort (2mg/kg at D1 and D5), the remaining patients of the second cohort received a single infusion of 2 mg/kg of XAV-19 at day 1. All patients also received therapy for COVID-19 according to standard of care (SOC) in the participating centres. This SOC included use of dexamethasone and other treatments per local practice and national guidelines at time of the study, which may include, not exclusively, antibiotics, antiviral treatment, immune therapies not based on antibodies administration and anticoagulants. Simple randomization was done using web-based simple (unstratified) allocation by trained clinical research staff. Study investigators, all research and analysis teams, and patients were masked to treatment allocation. The study medications were prepared and dispensed by the hospital pharmacy and presented as ready-to-use aqueous solutions in prelabelled infusion kits according to regulatory requirements. The study medication was then administered to the patients by the medical ward’ nurses, upon receipt.

### Procedures

Medical history and demographic data were collected at screening. Before dosing, patients underwent evaluation of vital signs, physical examination, assessments for pneumonia, blood haematology and chemistry as well as nasopharyngeal swab for SARS-CoV-2 RT-PCR. Patients also had a 12-lead electrocardiogram, and if clinically required chest x-rays and/or CT scan.

During and for the 2 hours after study treatment infusion, vital signs (temperature, respiratory rate, heart rate, systolic and diastolic blood pressures, and oxygen saturation) were monitored every 30 minutes, and particular attention was paid to occurrence of hypersensitivity or infusion-related reactions. Patients were followed for 2 months after the first infusion, with clinical assessment, including respiratory status and 7-point ordinal scale, daily during hospitalization, at days 8, 15, 29 and 60. Blood haematology and chemistry were collected at each visit. Samples to measure drug serum concentrations were collected in all patients at days 1 (pre-dose, post-dose), 3, 5 (pre-dose and post-dose, if appropriate), 8, 15, and 29. Drug concentration was assessed at Charles River Laboratories, Evreux, France, with a validated swine IgG-specific sandwich ELISA, presenting a lower limit of quantification of approximately 50 ng/mL in human serum. The use of concomitant medications was recorded throughout the study. Patients were also regularly assessed for adverse events and possible relation to the investigational medicinal product.

### Outcomes

The two primary outcomes were the pharmacokinetic measurement of the serum concentration of XAV-19 at day 8 and tolerability over 29 days, comparing the XAV-19 treated participants and placebo group. Pharmacokinetic parameters of XAV-19 that were evaluated were serum drug concentrations determined immediately at the end of infusion, maximum serum concentration (C_max_); time to maximum serum concentration (T_max_); area under the serum concentration–time curve extrapolated to infinity (AUC0–∞), which was calculated using the formula AUC_0–day 29_ (area under the serum concentration–time curve measured to the concentration at day 29 using the trapezoidal rule) and serum concentration at day 29/Ke (Ke is the apparent first-order terminal rate constant calculated from a semi-log plot of the serum concentration versus time curve); terminal half-life (t_1/2_) as determined by quotient 0.693/Ke; clearance, as determined by quotient dose/AUC0-∞ and volume of distribution, as determined by quotient clerarance/Ke.

Safety parameters were evaluated over 29 days by the onset of all adverse events suspected to be related to XAV-19 and incidence of serious adverse events, of treatment-related adverse events leading to discontinuation of study drug, of hypersensitivity reactions and infusion-related reactions.

Secondary and additional outcomes included pharmacokinetics of XAV-19 up to day 29 and clinical outcome (ICU transfer, 7-point ordinal scale, length of hospital stay). In an exploratory analysis, SARS-CoV-2 nasopharyngeal viral loads changes were assessed in an exploratory sub-cohort of patients treated by XAV-19 in comparison to the placebo group. Nasopharyngeal swabs were collected in 3mL of viral transport medium (Yocon, China). RNA extraction was done with EZ1 DSP virus kit (EZ1,Qiagen) according to manufacturer’s instructions. Viral nucleic acids were detected using a multiplex SARS-CoV-2 RT-qPCR, using *RdRp-IP2* and *RDRP IP4* adapted from Charité Protocol and National Reference Centre for Respiratory Viruses Institute Pasteur [12,13] When a sample was positive, quantification of the number of RNA copies was done by RT-qPCR using a specific *in vitro* transcribed RNA, according to a scale ranging from 2.2 to 10 log_10_ c/mL.

### Statistical analysis

The safety population included all subjects randomized into the study who received at least one dose of study drug. The intent-to-treat exposed (ITT-E) population was defined as all subjects who met study criteria and were randomized into the study with documented evidence of having received at least one dose of randomized treatment and at least one post-baseline measurement of serum XAV-19 titers. The per-protocol population was defined as all subjects included in the ITT-E population excluding those who had at least one major protocol deviation. Placebo patients were pooled for the purpose of analysis. The PK concentration population included all subjects who completed the study drug schedule. Categorical variables were summarized by percent and comparisons were assessed using an exact Fisher test. Continuous variables were summarized by means and standard error, and medians and interquartile range, comparisons between groups were done using a Kruskal-Wallis test. The primary endpoint was evaluated for ITT-E population by a Kruskal-Wallis test between placebo patients and treated patients. The number of adverse events was compared between groups of patients using an exact Fisher test. Comparisons of pharmacokinetic parameters were conducted using Kruskal-Wallis test. The relationship between AUC0–∞ and cumulative administered dose of XAV 19 was explored by a linear regression model. Groups of treatment were described according to variables in of the secondary endpoints. A p-value < 0.05 was defined as significant.

## RESULTS

Between Aug 30^th^, 2020 and Dec 7^th^, 2020, 18 patients with COVID-19-related moderate pneumonia were randomized, one withdrew consent before day 5 second infusion (cohort 1) and 17 completed all study visits (**Supplemental Figure 1**). Hence the safety population is constituted of 18 patients, whereas the ITT-E population was constituted of 17 patients. Demographics and baseline characteristics of the 17 patients were similar across groups (**Table 1**). Of the 17 analyzed patients, 12 were treated with XAV-19 (0.5 mg/kg at days 1 and 5 n=6, 2 mg/kg at days 1 and 5 n=1, 2 mg/kg at day 1 n=5) and five with placebo, 11 were males (64.7%), median age was 71 years [interquartile range (IQR) 51-75], and median body mass index (BMI) was 27.4 [25.3–31.2] kg/m^2^. The majority (14/17) of patients had at least one comorbidity, including three with immunodeficiency disease (**Table 1**). At screening, all patients were hospitalized, requiring supplemental oxygen by nasal prongs or mask with low oxygen delivery (WHO scale 4). Associated COVID-19 therapies were steroids in all patients and remdesivir in 8 (47%) patients. The median time from symptom onset to the initial infusion of investigational drug was 8 days [IQR 6-9].

**Table 1.** Baseline characteristics of patients and clinical outcome.

| Characteristic | XAV-19 (n=12) |  |  | Placebo (n=5) | Total (n=17) |
| --- | --- | --- | --- | --- | --- |
|  | 0.5 mg/kg<br>D1-D5<br>(n=6) | 2 mg/kg<br>D1-D5<br>(n=1) | 2 mg/kg<br>D1<br>(n=5) |  |  |
| Age, years, median (IQR) | 68 (49.8; 77.3) | 75 | 74 (68; 76) | 63 (51; 71) | 71 (51; 75) |
| Males, *n* (%) | 3 (50) | 1 | 4 (80) | 3 (60) | 11 (64.7) |
| BMI, kg/m<sup>2</sup>, median (IQR) | 25.1 (23.6; 29.8) | 39.2 | 27.4 (27.4; 28.4) | 29.3 (26.1; 29.8) | 27.4 (25.3; 31.2) |
| Chronic underlying disease, *n* (%) |  |  |  |  |  |
| Asthma | 2 (33.3) | 0 | 0 | 1 (20) | 3 (17.6) |
| Cardiovascular/cerebrovascular disease | 1 (16.7) | 1 | 1 (20) | 2 (40) | 5 (29.4) |
| Chronic kidney disease | 1 (16.7) | 0 | 0 | 0 | 1 (5.9) |
| Diabetes | 0 | 0 | 1 (20) | 1 (20) | 2 (11.8) |
| High blood pressure | 4 (66.7) | 1 | 2 (40) | 1 (20) | 8 (47.1) |
| Obesity | 1 (16.7) | 0 | 1 (20) | 1 (20) | 3 (17.6) |
| Solid organ transplant | 1 (16.7) | 0 | 0 | 0 | 1 (5.9) |
| Solid tumor | 1 (16.7) | 0 | 1 (20) | 0 | 2 (11.8) |
| HIV infection | 1 (16·7) | 0 | 0 | 0 | 1 (5·9) |
| Duration of symptoms before first infusion, median (IQR) | 7·5 (5·3; 9·8) | 5 | 8 (6; 9) | 8 (8; 8) | 8 (6; 9) |
| Symptoms at infusion, *n* (%) |  |  |  |  |  |
| Hospitalized, without oxygen supplementation | 1 (16·7) | 0 | 0 | 1 (20) | 2 (11·8) |
| Hospitalized, low oxygen delivery | 5 (83·3) | 1 | 4 (80) | 4 (80) | 14 (82·4) |
| Hospitalized, on NIV or high flow | 0 | 0 | 1 (20) | 0 | 1 (5·9) |
| Respiratory rate (breath/min), median (IQR) | 27 (22·5; 31·5) | 26 | 22 (20; 28) | 24 (24; 28) | 24 (22; 30) |
| SpO<sub>2</sub>, median (IQR) | 94 (93·3; 94·8) | 96 | 92 (91; 94) | 94 (93; 94) | 94 (92; 92) |
| NewS 2, median (IQR) | 6·5 (6; 7·8) | 6 | 7 (5; 8) | 6 (6; 6) | 6 (5; 7) |
| Concomitant medication for COVID-19 |  |  |  |  |  |
| Antibiotics | 4 (66·7) | 0 | 2 (40) | 2 (40) | 8 (47·1) |
| Steroids | 6 (100) | 1 | 5 (100) | 5 (100) | 17 (100) |
| Remdesivir | 2 (33·3) | 1 | 1 (20) | 4 (80) | 8 (47·1) |
| Anticoagulant | 5 (83·3) | 1 | 5 (100) | 5 (100) | 16 (94·1) |
| Laboratory parameters at D1, median (IQR) |  |  |  |  |  |
| White blood cells/mm<sup>3</sup> | 7·9 (7·6; 10·3) | 4·2 | 7·2 (6·9; 7·5) | 6·5 (6·2; 6·7) | 7·1 (6·5; 7·9) |
| Haemoglobin | 11·8 (11; 12·5) | 12·9 | 12·7 (11·9; 13·4) | 14·1 (13·2; 15) | 12·4 (11·3; 13·3) |
| Platelets /mm<sup>3</sup> | 314 (238·8 ;407·5) | 163 | 257 (237; 276) | 244.5 (191; 297) | 261 (172; 350) |
| Creatinine <sup>a</sup> | 85 (76·3 ;92·8) | 86 | 65·5 (64; 67) | 76 (71·5; 80·5) | 84 (67; 86) |
| ICU admission, *n* (%) | 0 | 0 | 2 (40) | 1 (20) | 3 (17·6) |
| Hospital length of stay, days, median (IQR) | 5·5 (5 ;11·3) | 10 | 11 (7; 12) | 7 (7; 12) | 7 (6; 12) |
| 60-day mortality, *n* (%) | 1 (14·3) | 0 | 0 | 0 | 1 (5·6) |

<sup>a</sup> missing data: 0.5 mg/kg group at day 1 and 5 (n=2), 2 mg/kg group at D1 (n=3), placebo group (n=3).

Pharmacokinetic parameters of XAV-19 are presented **in Table 2**. XAV-19 serum concentrations (median [range]) at day 8 were 6.4 μg/mL [2.8-11.9] in patients who received 0.5 mg/kg of XAV-19 at days 1 and 5, 20.3 μg/mL (12.0-22.7) for those who received 2 mg/kg at day 1, 47.2 for for the one who received 2 mg/kg at days 1 and 5 (p=0.012 between groups) (**Table 2**). Serum concentrations were above 29 μg/mL in all patients at day 1 post-infusion of 2 mg/kg of XAV-19 (**Table 2**). Based on a target neutralization threshold of 10 μg/mL of XAV-19, all patients treated with 2 mg/kg (1 or 2 infusions) had serum concentrations above the target at day 8. A proportional relationship between cumulative administered dose and AUC0–∞ was found suggesting a linear pharmacokinetic for XAV-19 between these doses (**Figure 1**). According to this linearity, no significant difference between t_1/2_ values was observed between the groups of patients. The half-life [median; range; n =12] for all XAV-19 doses was 13·0 (0.7-19.4) days, which allowed to maintain serum concentration of XAV-19 above the previously defined target serum level of 10 μg/mL (two fold the 100% neutralization activity in vitro) during 15 days after a single 2 mg/kg administration for most patients (**Figure 2**) [9]. After a single infusion of the 2 mg/kg dose, the volume of distribution (L) and the clearance (L/h) (median; range; n=5) were respectively 4.9 (4.0-8.7) and 0.015 (0.010-0.021).

**Figure 1.**
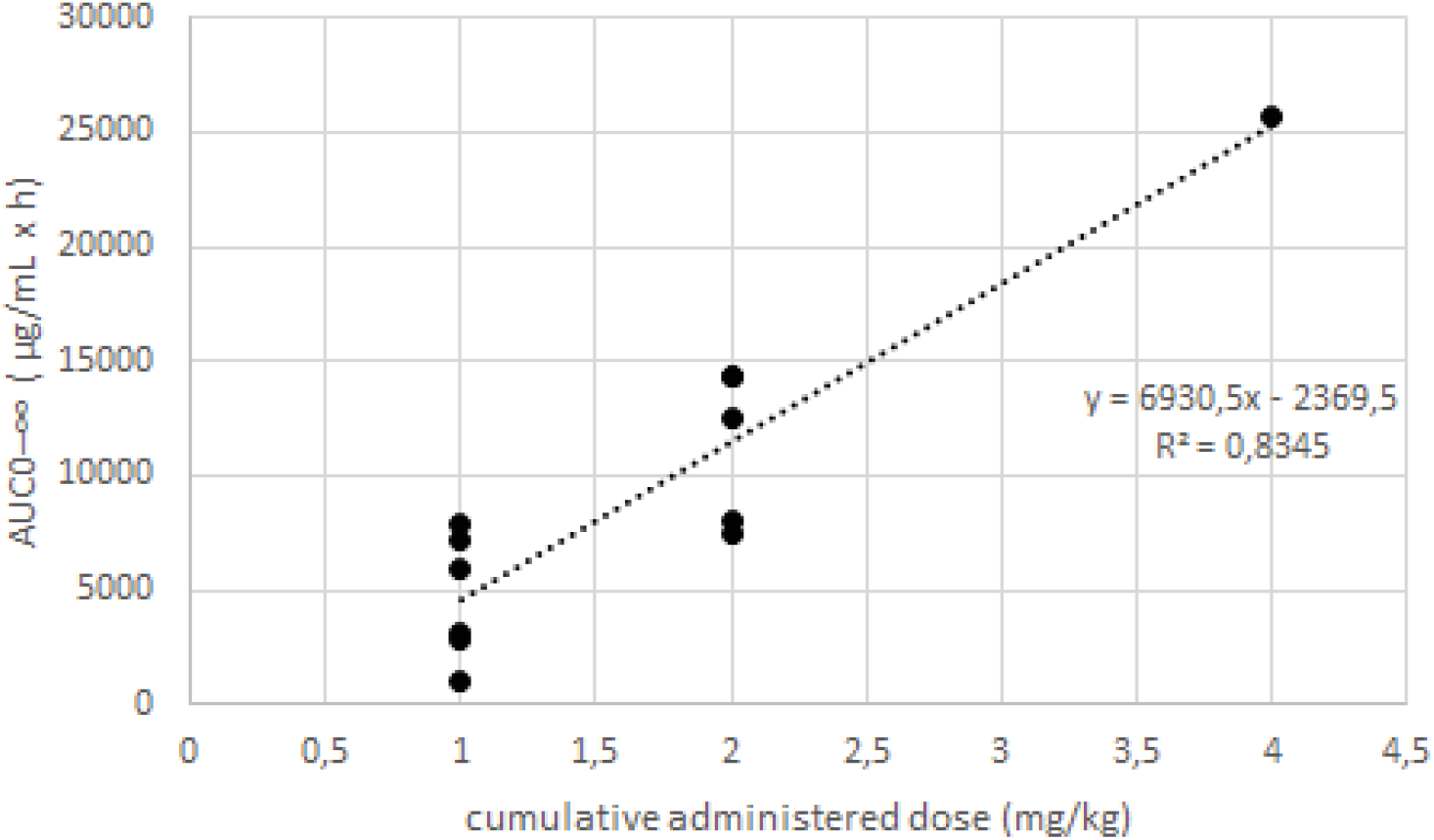
Relationship between AUC0–∞ and cumulative administered dose of XAV-19.

**Figure 2.**
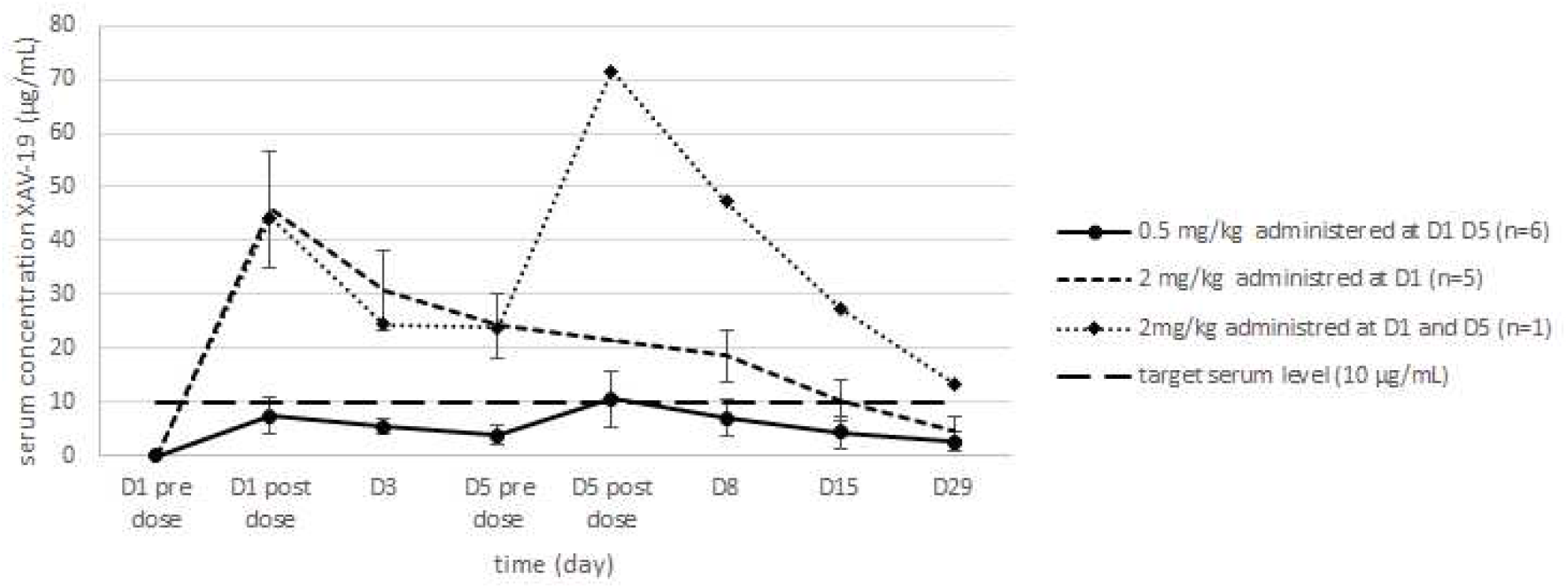
Time course of XAV-19 mean (+/- standard deviation) serum concentration in the groups of patients. D = day. D = day

**Table 2.** XAV-19 pharmacokinetic-parameter estimates according to dose and schedule of administration.

| Variable | XAV-19 (n=12) |  |  | *P value* |
| --- | --- | --- | --- | --- |
|  | 0.5 mg/kg | 2 mg/kg | 2 mg/kg |  |
|  | D1 and D5<br>(n=6) | D1 and D5<br>(n=1) | D1<br>(n=5) |  |
| Concentration post day 1 infusion,<br>median (min-max) | 6.90 (3.29-10.9) | 47.35 (29.10-55) * |  | 0.0022 |
| Serum concentration at D8<br>(µg/mL), median (min-max) | 6.4 (2.8-11.9) | 47.2 | 20.3 (12.0-22.7) | 0.012 |
| AUC<sub>0-∞</sub> (µg/mL x h), median<br>(min-max) | 4,564 (987-7,832) | 2,5638 | 12.519 (7.515-14.314) | 0.018 |
| C<sub>max</sub> (µg/mL), median (min-max) | 9.1 (5.2-18.1) | 71.5 | 50.4 (29.1-55.0) | 0.012 |
| T<sub>max</sub> | Day 5 post dose | Day 5 post dose | Day 1 post dose |  |
| t<sub>1/2</sub> (days), median (min-max) | 14.5 (0.7-19.4) | 11.88 | 11.4 (5.5-13.9) | 0.211 |

\* values for all Day 1 post-infusion measures at the dose of 2 mg/kg (n = 6).
AUC, Area under the serum concentration–time Curve; C<sub>max</sub>, maximum serum concentration; T<sub>max</sub>, time that a drug is present at the maximum concentration in serum; t<sub>1/2</sub>, half-life.

A summary of the adverse events is presented **in Table 3**. XAV-19 was well tolerated, with no differences in adverse events nature and rate between placebo and XAV-19 group and no serious adverse events related to the study drug. Only two patients experienced drug-related grade 1 adverse event (one had lymphadenopathy in the placebo group, one had localized transient ecchymotic rash at day 15 in the 0·5 mg/kg group). Severe adverse events occurred in three patients in the placebo group with two grade 3 events (worsening asthenia and increased dyspnoea) and one grade 4 event (respiratory distress), and in five patients in the XAV-19 group with two grade 4 events (worsening respiratory distress) and three grade 3 events (respiratory failure and two hepatobiliary disorders) (**Supplemental Tables 1 and 2**). No hypersensitivity or infusion-related reactions were reported during treatment, and there were no treatment discontinuations due to adverse events. There were no consistent or clinically significant changes in haematology, clinical chemistry, vital signs, or urinalysis values. Of the 17 patients included in the ITT-E population, three (17.6%) developed respiratory failure requiring high flow ventilation, one (20%) in the placebo group and two (16.6%) in patients who received XAV-19. At day 8, 11/17 patients were free of oxygen, 3/5 in the placebo group and 8/12 in the XAV-19 group. At last follow-up visit, on day 60, one patient had died at day 58, from complications of comorbidities in the 0.5 mg/kg group and all other patients showed clinical improvement and had been discharged from the hospital (Figure 3). Longitudinal assessment of quantitative viral load in nasopharyngeal swab was available in four patients in the 2 m/kg XAV-19 groups and in five patients in the placebo group, suggesting a trend to faster clearance of SARS-CoV-2 with XAV-19 (**Figure 4**).

**Table 3.** Adverse events (safety analysis population) over 29 days, numbers of patients (%) with at least 1 event in class.

| Adverse event | XAV-19 (n=12) |  |  | Placebo (N=5) |
| --- | --- | --- | --- | --- |
|  | 0.5 mg/kg | 2 mg/kg | 2 mg/kg |  |
|  | D1 and D5 | D1 and D5 | D1 |  |
|  | (n=7) | (n=1) | (n=5) |  |
| Number of patients (percent) |  |  |  |  |
| Adverse events |  |  |  |  |
| Any (per patient), Mean (SD) | 3.7 (2.7) | 5 | 3 (2.6) | 5.6 (4) |
| Grade 1 or 2 | 7 (100) | 1 (100) | 4 (80) | 5 (100) |
| Grade 3 or 4 | 2 (28.6) | 0 | 2 (40) | 3 (60) |
| Adverse events related to study drug | 1 | 0 | 0 | 1 |
| Number of patients (%) with $\geq 1$ adverse events by class according to MedDRA (System Organ Class) | | | | |
| Blood and lymphatic system disorders | 3 (42.9) | 0 | 0 | 1 (20) |
| Cardiac disorders | 0 | 1 (100) | 0 | 0 |
| Gastrointestinal disorders | 1 (14.3) | 0 | 0 | 2 (40) |
| General disorders and administration site conditions | 1 (14.3) | 1 (100) | 1 (20) | 3 (60) |
| Hepatobiliary disorders | 1 (14.3) | 1 (100) | 2 (40) | 3 (60) |
| Infections and infestations | 3 (42.9) | 0 | 0 | 0 |
| Injury, poisoning and procedural complications | 1 (14.3) | 0 | 0 | 0 |
| Musculoskeletal and connective tissue disorders | 2 (28.6) | 0 | 0 | 1 (20) |
| Neurological/ psychiatric system disorders | 1 (14.3) | 0 | 1 (20) | 2 (40) |
| Renal and urinary disorders | 4 (57.1) | 0 | 1 (20) | 0 |
| Respiratory, thoracic and mediastinal disorders | 1 (14.3) | 1 (100) | 3 (60) | 3 (60) |
| Skin and subcutaneous tissue disorders | 1 (14.3) | 0 | 0 | 2 (40) |

**Figure 3.**
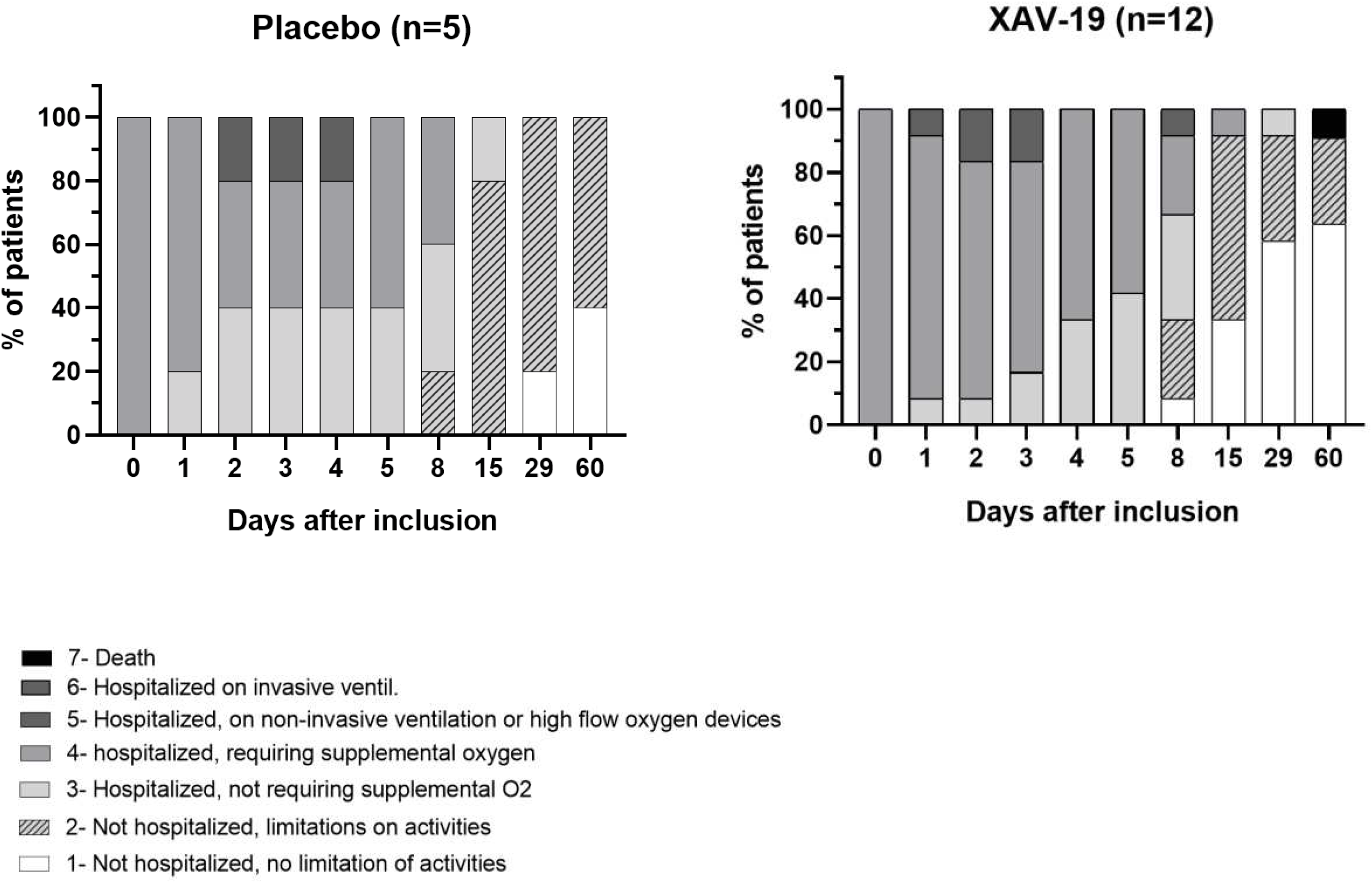
Clinical Status According to 7-Category WHO Ordinal Scale between screening (day 0) and last-follow-up (day 60) in placebo (n = 5) and XAV-19 (n = 12) groups.

**Figure 4.**
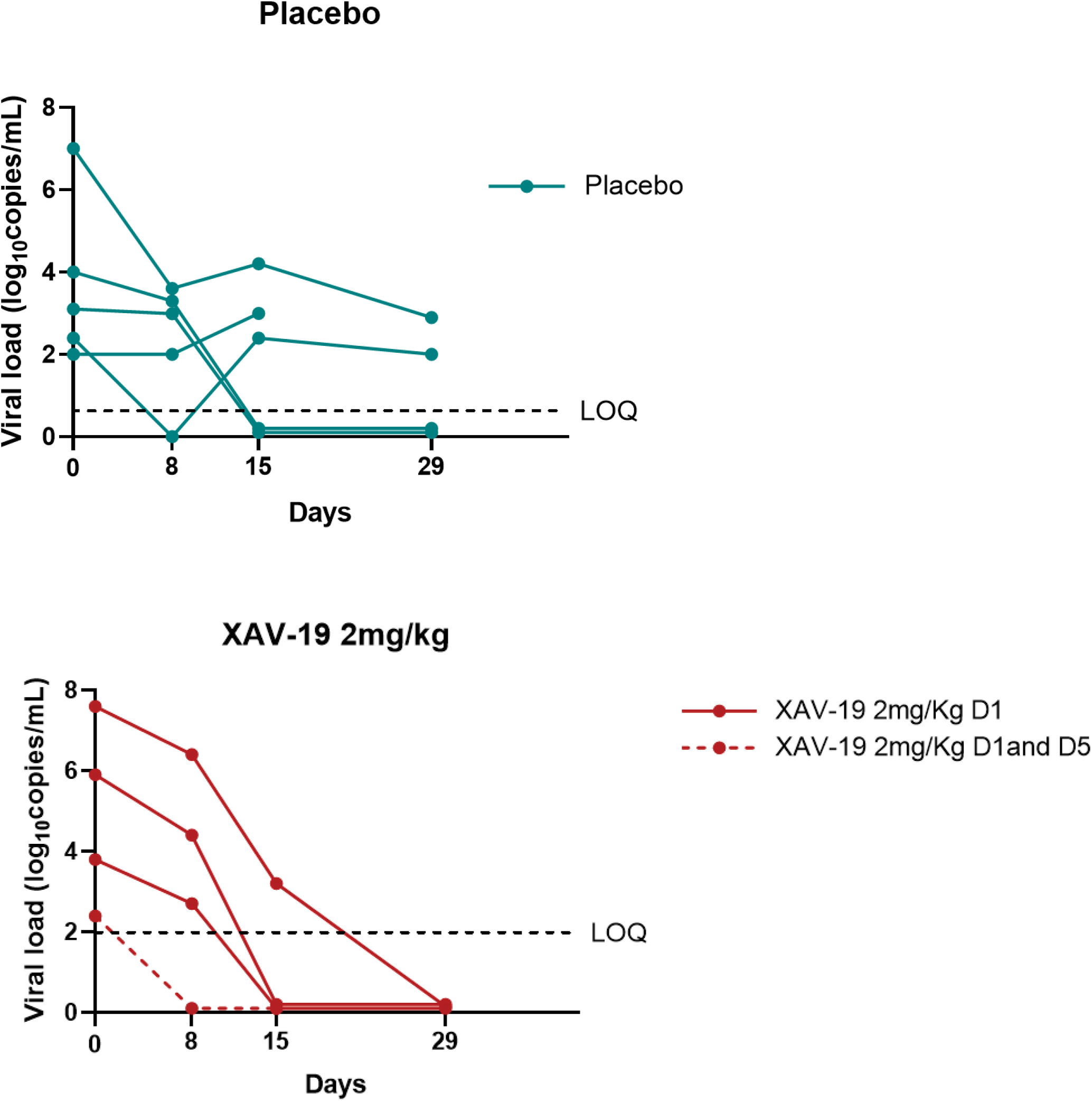
Nasopharyngeal viral load over time in placebo (n = 5) and XAV-19 2mg/kg (n = 4) groups. D = day, LOD= limit of detection (2.2 log_10_ copies/mL), 0 = no signal detected

## DISCUSSION

This study was a randomized, double-blind, placebo-controlled, multicentre, phase IIa trial evaluating the optimal dose and the safety of XAV-19, a swine-derived glyco-humanized polyclonal SARS-CoV-2-neutralizing antibody, in patients admitted to hospital with COVID-19-related moderate pneumonia. The results of this first-in-human trial have shown that a single infusion of XAV-19 at 2 mg/kg maintained serum concentration of XAV19 well above the predicted neutralization target concentration of 10μg/mL during at least 8 days post-infusion [9]. The median elimination half-life for XAV-19 2 mg/kg was estimated at 11.4 days, which provides rationale for a single infusion to maintain in vivo neutralizing activity for at least 8 days. Our data also suggest that XAV-19 might enhance viral clearance, even in patients admitted to hospital after on average one week of onset of SARS-CoV-2 symptoms, although our analysis was exploratory and performed on a limited number of patients. Studies have shown that symptom onset is not a sufficient predictor of level of viral load in respiratory secretions, as high viral loads might persist for 2 weeks, nor of individual innate immune response or ability of this immune response to control ongoing viral replication[14–16]. Then, although maximal benefit of any antiviral therapy, either specific antiviral or neutralizing antibodies, is expected when treatment is started earlier in the illness, benefit could also persist in patients treated with longer duration of symptoms, as demonstrated with remdesevir[6].

XAV-19 maintains neutralization activity against the most predominant SARS-CoV-2 variants, including the initial Wuhan strain and the 501Y.V1 and 501Y.V2 variants, while some anti-SARS-CoV-2 monoclonal antibodies exhibit reduced activity against the new variants[11]. Indeed, bamlanivimab has slight loss of activity against the B.1.1.7 variant, while both bamlanivimab and etesivimab have complete loss of activity against the B.1.351 variant.; casivirimab has large loss of activity against the B.1.351 variant and imdevimab retains similar activity against all strains. Interim results of recent trials have suggested that among non-hospitalized patients with mild to moderate COVID-19 illness not requiring oxygen supplementation, treatment with a combination of 2 anti-spike neutralizing monoclonal antibodies was associated with antiviral effect. To what extent such effect translates in clinical benefit by reducing worsening and requirement for hospitalization is being assessed in phase III trials, especially in high-risk patients[17–19]. As new strains of SARS-CoV-2 of epidemiological importance might continue to emerge, it will be necessary to carefully determine both in vitro and in vivo the neutralizing activity of cocktails of monoclonal antibodies and of XAV-19. Polyclonal antibodies offer the advantages over monoclonal antibodies to cover the different epitopes of the target antigen and to mimic natural responses to the antigen, with in addition a lower cost.

XAV-19 was well tolerated and no major safety issues or dose-related trends were identified. The clinical outcome of COVID-19 was not different in both groups, but numbers were too small in this phase IIa study to see any trend. Rate of worsening of respiratory failure, based on WHO scale, was within expected range based on characteristics of enrolled population, and the death observed at day 59 was unrelated to study drug or COVID-19 [20,21]. No immediate hypersensitivity reactions or infusion-related reactions were reported in our study contrasting to reports and warning with cocktail of anti-spike monoclonal antibodies [19].

An important limitation of this phase IIa portion of our trial is the small sample size, that did not allow to determine if XAV-19 was associated with improved outcomes. This question will be rigorously tested in the analysis of the phase III part of this ongoing trial. Even if the number of patients who received XAV-19 was low, there were no safety concerns. Second, higher doses of XAV-19 were not explored. However, the 2 mg/kg dose achieved sustained active concentrations.

In conclusion, XAV-19 was well tolerated in patients admitted to hospital for COVID-related moderate pneumonia requiring low-flow oxygen supplementation. The pharmacokinetic results of a single infusion of 2 mg/kg suggest that this dose has the potential to successfully block viral diffusion in humans and supports the selection of this regimen for the ongoing multicentre randomized (1:1), double-blind, placebo-controlled phase III trial (ClinicalTrial.gov, NCT04453384) involving 400 patients.

## Data Availability

The data analysed and presented in this study are available from the corresponding author on reasonable request, providing the request meets local ethical and research governance criteria after publication. Data collected during the study may be processed electronically, in accordance with the requirements of the CNIL (compliance with reference methodology MR001).

## Authors’ contributions

BG and FR conceptualised the study. BG, BV, RJ, SB, OD, AO, BL, OD, and FR were involved in finalizing the protocol of the study. BG, FR, LB, and AO supervised the study. BG, KL, VD, FA and FR were involved in the clinical care of the patients. AC, AJ, LB were involved in data curation. ED, BV, VF MAV, RJ, SB, RD, ALT, AC and AJ did formal analysis of data. LF was responsible of central depository and distribution of study drugs. BG and FR prepared the original draft of the manuscript. All authors were involved in writing, reviewing, and editing of the manuscript.

## Data sharing

The data analysed and presented in this study are available from the corresponding author on reasonable request, providing the request meets local ethical and research governance criteria after publication. Data collected during the study may be processed electronically, in accordance with the requirements of the CNIL (compliance with reference methodology MR001). The study protocol is provided in the appendix.

### Funding

This work was funded by Nantes University Hospital Research Department with support from Public Investment Bank (BPI France) in the framework of the “Investment for the Future” programme (Programme d’Investissements d’Avenir) and Xenothera. The funder of the study had a role in study design, data collection, data analysis. The corresponding author and co-authors interpreted the data, wrote the report had full access to all the data in the study and had final responsibility for the decision to submit for publication.

## Acknowledgements

We thank clinical and research teams, as well as pharmacists and virologists at all the participating clinical centres, who contributed to the management of the study patients for their commitment to providing optimal patient care.

## *POLYCOR study group

### Trial development team

Laetitia Berly, Sophie Brouard, Odile Duvaux, Laurent Flet, Benjamin Gaborit, Régis Josien, Alexandra Jobert, Aurélie Le Thuaut, Anne Omnès, François Raffi, Emilie Rebouilleau, Laurent Vacher, Bernard Vanhove, Marie-Anne Vibet.

### Trial management team

Anne Chiffoleau, Laetitia Berly, Benjamin Gaborit, Marion Gautier, François Raffi, Joseph Herault, Alexandra Jobert, Aurélie Le Thuaut, Anne Omnès, Ludivine Perrier, Emily Rebouilleau, Sandrine Renaud, Marie-Anne Vibet.

### Trial Steering Comité

Florence Ader, Odile Duvaux, Virginie Ferre, Benjamin Gaborit, Karine Lacombe, Anne Omnès, François Raffi, Bernard Vanhove, Eric Vicaut

### Trial Independent data monitoring committee

Bruno Hoen, Laura Richert, Caroline Solas, Astrid Vabret

### Investigators

<u>Angers :</u> Marc-Antoine Custaud, Valérie Daniel, Vincent Dubee, Rafaël Mahieu ; <u>Nantes :</u> Cécile Braudeau, Marie Chauveau, Eric Dailly, Colin Deschanvres, Laurent Flet, Benjamin Gaborit, Matthieu Gregoire, Anne-sophie Lecomte, Maëva Lefebvre, Pascale Morineau Le Houssine, François Raffi, Martine Tching-Sin. ; <u>Lyon :</u> Florence Ader, Agathe Becker, Pierre Chauvelot, Anne Conrad, Tristan Ferry, Julianne Oddone, Thomas Perpoint, Cécile Pouderoux, Sandrine Roux, Claire Triffault-Filit, Florent Valour ; Paris <u>Saint Antoine :</u> Diane Bollens, Thibault Chiarabini, Anne Daguenel-Nguyen, Emmanuelle Gras, Patrick Ingiliz, Karine Lacombe, Bénédicte Lefebvre, Laura Levi, Zineb Ouazene, Jérôme Pacanowski, Laure Surgers, Nadia Valin.

## Supplemental material

**Supplemental Figure 1.**
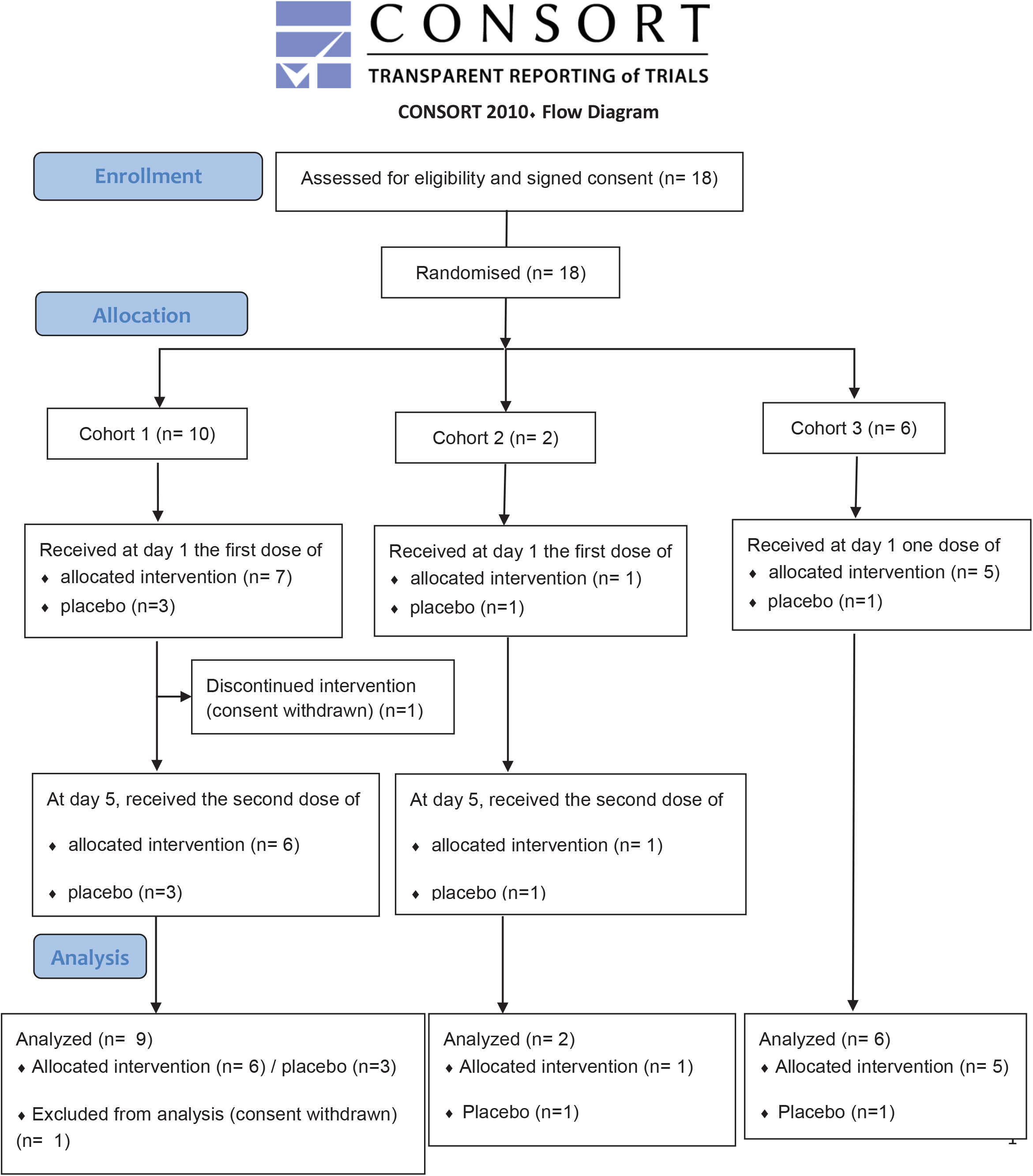
Flow diagram

**Supplemental Table 1.**
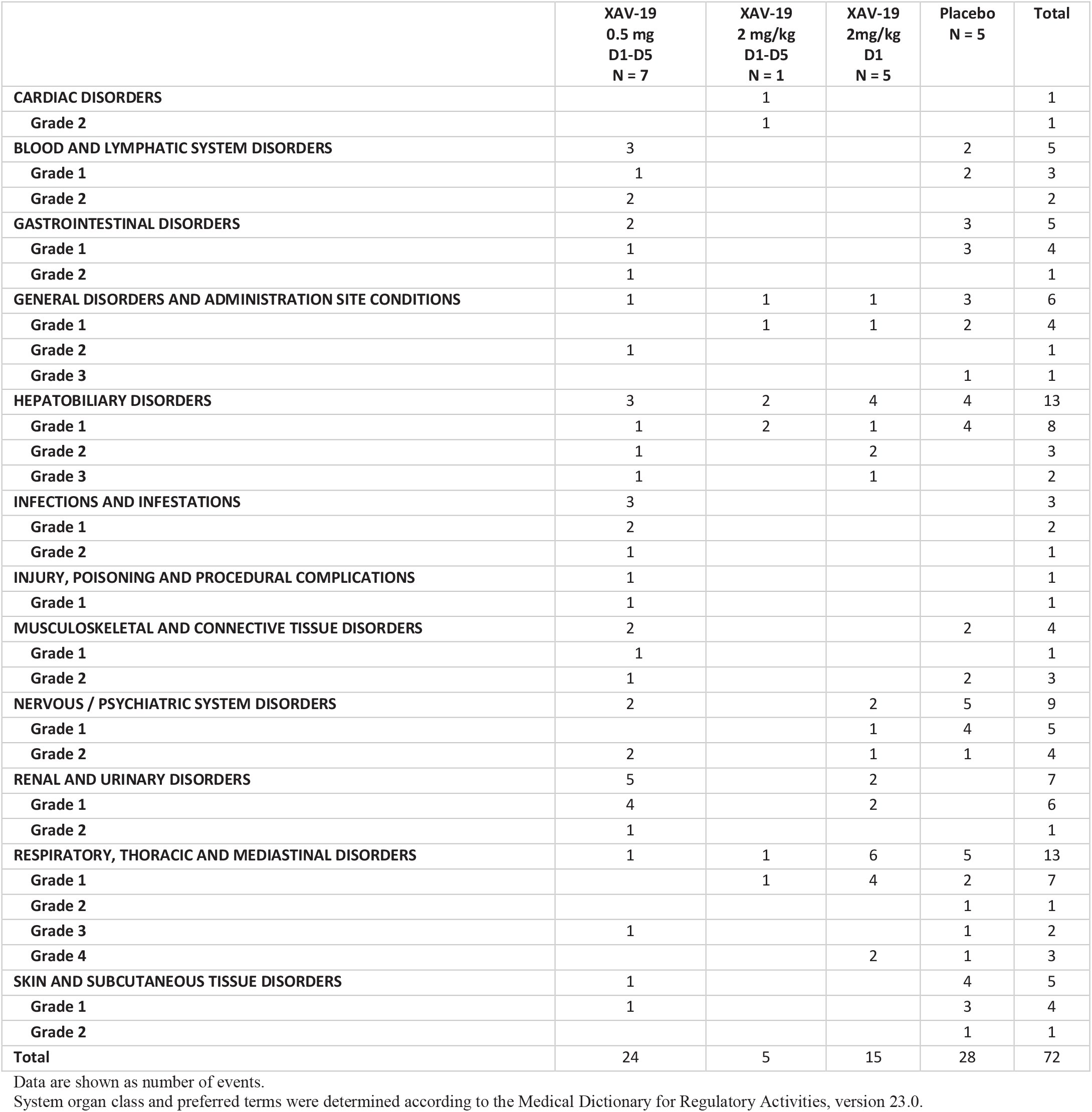
Adverse events by system organ class and grade occurring in either treatment group to day 29 (safety population).

**Supplemental Table 2.**
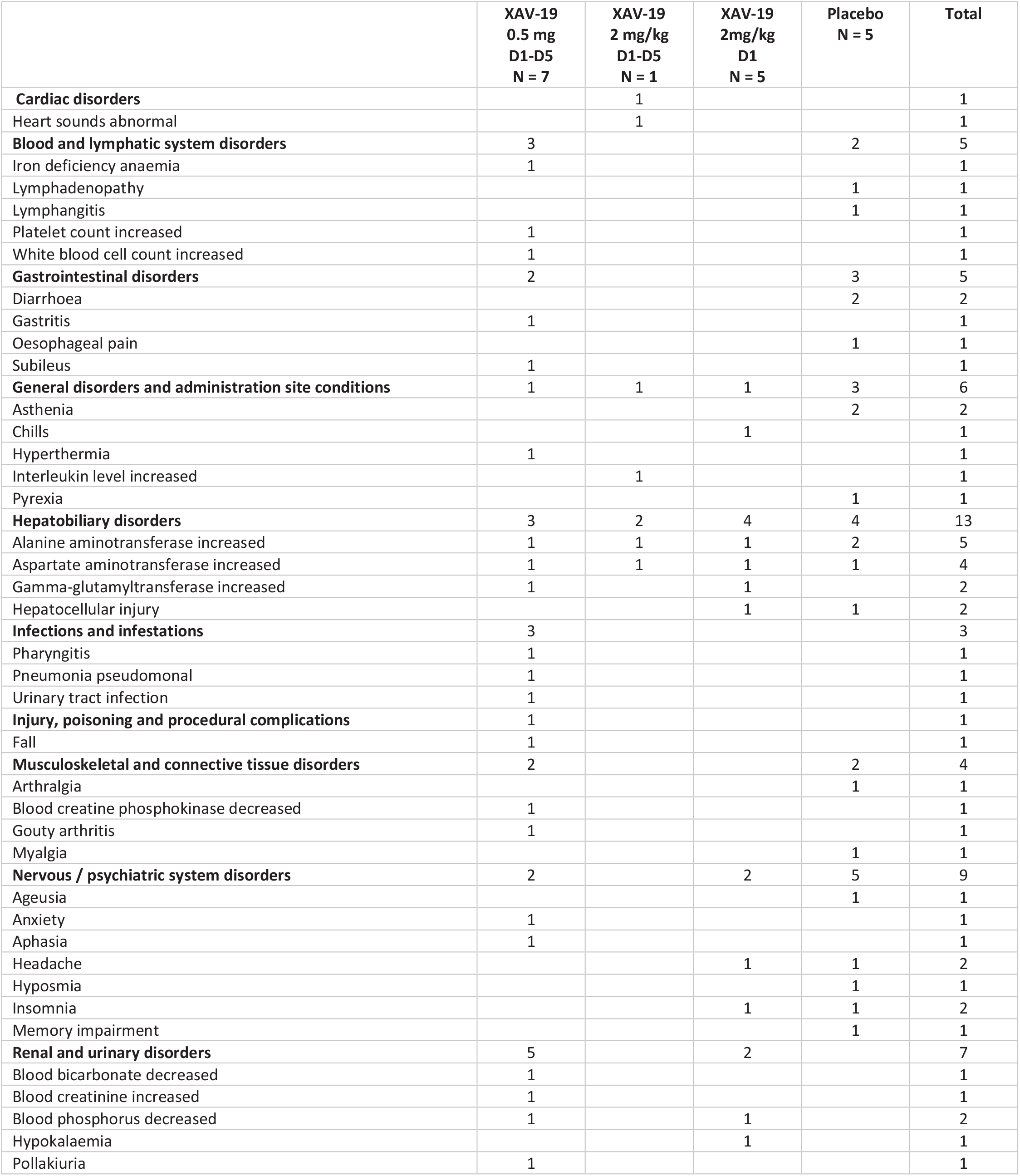
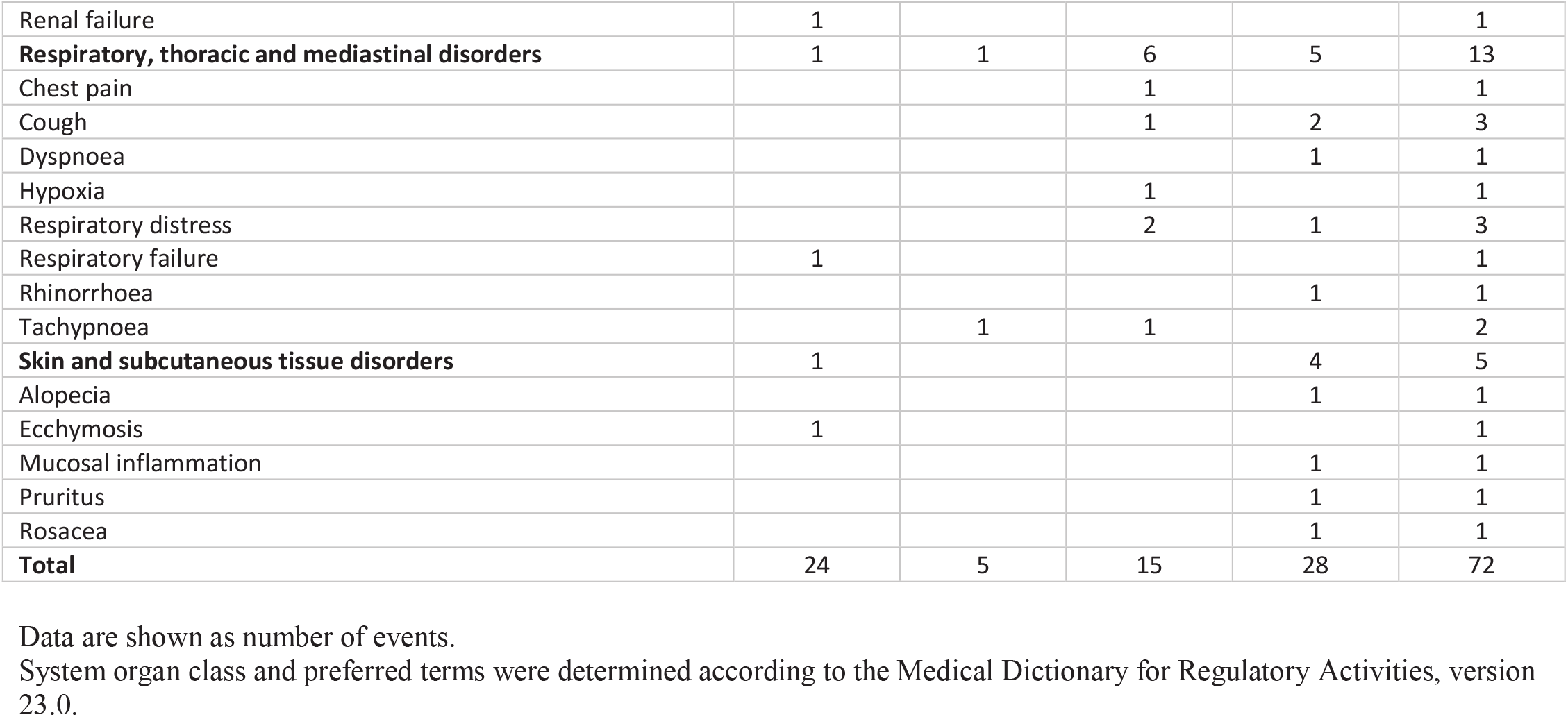
Adverse events by system organ class and preferred term occurring in either treatment group to day 29 (safety population).

